# Genome-wide colocalization of body fat distribution GWAS and subcutaneous adipose eQTLs identifies SNX10, DGKQ, and CBX3 as candidate causal genes for cardiometabolic disease

**DOI:** 10.64898/2026.06.13.26355580

**Authors:** Muhammad Shahzad Iqbal

## Abstract

**Background:** Genome-wide association studies (GWAS) have identified hundreds of loci associated with body fat distribution, yet the causal genes and regulatory mechanisms through which these variants exert their effects remain largely unknown. Expression quantitative trait locus (eQTL) colocalization provides a powerful framework for identifying genes whose expression is genetically coregulated with complex traits.

**Methods:** We performed a genome-wide colocalization analysis integrating waist-hip ratio adjusted for body mass index (WHRadjBMI) GWAS summary statistics from 694,649 individuals with subcutaneous adipose tissue eQTLs from the Genotype-Tissue Expression (GTEx) Project v8 (N = 581 donors). GWAS coordinates were lifted from GRCh37 to GRCh38 to enable direct alignment with GTEx data. We incorporated CAVIAR fine-mapping results to overcome the limitation of FDR-significant eQTL filtering. Colocalization was assessed using the approximate Bayes factor framework (coloc.abf) across 335 independent genome-wide significant loci.

**Results:** Of 2,897 locus-gene pairs tested, 489 (16.9%) showed strong colocalization (PP.H4 > 0.8) and 618 (21.3%) showed moderate evidence (PP.H4 > 0.5). The strongest colocalization was observed for SNX10 (sorting nexin 10; PP.H4 = 1.000), a recently characterized regulator of adipocyte differentiation and female-specific diet-induced obesity. Other top hits included DGKQ (diacylglycerol kinase theta; PP.H4 = 0.9999999), an emerging pharmacological target for insulin resistance, and CBX3 (chromobox 3; PP.H4 = 0.9999974), an epigenetic regulator linked to cardiovascular disease. Established adiposity genes including GRB14 (PP.H4 = 0.681) and KLF14 (PP.H4 = 0.590) were recovered, validating our approach. Several loci exhibited extensive allelic heterogeneity, with 50 genes colocalizing at a single chromosome 3 locus.

**Conclusions:** Our analysis provides a comprehensive map of adipose tissue gene regulatory mechanisms underlying genetic risk for body fat distribution. The identification of SNX10, DGKQ, and CBX3 as high-confidence candidate causal genes advances the translation of GWAS associations into mechanistic understanding and therapeutic targets for obesity-related cardiometabolic disease.

## 1. Introduction

Obesity is a global health crisis affecting more than one in three adults worldwide, with profound consequences for cardiometabolic disease including type 2 diabetes, cardiovascular disease, and certain cancer. While general adiposity, measured by body mass index (BMI), is a well-established risk factor, epidemiological evidence increasingly indicates that the location and distribution of excess fat—rather than its absolute amount—are more informative for predicting disease risk. Individuals with higher central adiposity, as assessed by waist-to-hip ratio (WHR), face elevated cardiometabolic risk independent of their BMI, whereas those with greater gluteal adiposity appear protected (Phelps et al., 2024). This body fat distribution phenotype is highly heritable, with twin-based estimates ranging from 30% to 60% and narrow-sense heritability reaching approximately 50% in women and 20% in men (Hansen et al., 2023).

The largest genome-wide association study (GWAS) meta-analysis of body fat distribution, conducted by the GIANT consortium and UK Biobank, identified 463 signals across 346 loci associated with WHR adjusted for BMI (WHRadjBMI) in 694,649 individuals of European ancestry (Pulit et al., 2019). Notably, approximately one-third of these signals exhibited sexual dimorphism, with stronger effects in women, and the top 5% of individuals carrying WHRadjBMI-increasing alleles were 1.62 times more likely to exceed metabolic syndrome thresholds. Despite this remarkable progress in variant discovery, the causal genes and regulatory mechanisms through which these GWAS signals exert their effects on adipose tissue biology remain largely unresolved (Hansen et al., 2023).

Expression quantitative trait locus (eQTL) colocalization has emerged as a powerful integrative framework for bridging this gap (GTEx Consortium, 2017). By testing whether the same causal variant drives both a complex trait association and local gene expression, colocalization distinguishes true regulatory mechanisms from coincidental LD [cite: web_search:17#3]. The Genotype-Tissue Expression (GTEx) Project has mapped eQTLs across 49 tissues, with subcutaneous adipose tissue representing a primary depot for energy storage and a key site of metabolic dysregulation (GTEx Consortium, 2017). However, standard colocalization approaches face practical challenges: GTEx significant eQTL files contain only FDR-significant associations, omitting the full LD landscape required for robust Bayes factor computation, and genome build mismatches between legacy GWAS (GRCh37) and GTEx (GRCh38) have historically hindered direct integration.

To address these limitations, we performed a genome-wide colocalization analysis integrating the largest WHRadjBMI GWAS with subcutaneous adipose eQTLs from GTEx v8. We employed liftOver to align GWAS coordinates to GRCh38, incorporated CAVIAR fine-mapping results to expand variant coverage beyond FDR-significant eQTLs, and used the approximate Bayes factor framework (Giambartolomei et al., 2014) to systematically test 335 genome-wide significant loci. Our analysis identifies 489 strong colocalizations, including novel candidate causal genes with established roles in adipocyte biology and emerging therapeutic relevance, providing a resource for translating GWAS associations into mechanistic understanding of body fat distribution and its cardiometabolic consequences.

## 2. Methods

### 2.1. GWAS Data

Summary statistics for waist-hip ratio adjusted for body mass index (WHRadjBMI) were obtained from the GIANT and UK Biobank meta-analysis (Pulit et al., 2019) conducted by Pulit et al. (2019). The dataset comprised 694,649 individuals of predominantly European ancestry. The original release was mapped to the GRCh37/hg19 genome build and restricted to HapMap2 variants (∼2.5 million SNPs). To enable integration with GTEx v8 eQTL data (GRCh38), GWAS coordinates were lifted from hg19 to hg38 using the UCSC liftOver tool (chain file: hg19ToHg38.over.chain.gz) (Hinrichs, 2006; Kent et al., 2002). Of 2,516,184 input SNPs, 2,515,080 (99.96%) were successfully mapped; 2,208 SNPs were unmapped and excluded. The lifted dataset contained 2,515,080 variants with the following columns: chromosome (CHR), position (POS), SNP identifier (rsID with alleles), tested allele, other allele, frequency of the tested allele, effect size (BETA), standard error (SE), P-value (P), sample size (N), and imputation quality score (INFO).

### 2.2. Quality Control of GWAS Data

Prior to colocalization analysis, GWAS variants were filtered to retain high-quality common variants: imputation INFO score > 0.8, minor allele frequency (MAF) between 1% and 99% (corresponding to Freq_Tested_Allele > 0.01 and < 0.99), positive standard error (SE > 0), non-missing BETA, and valid P-values (0 < P <= 1). After QC, 2,482,488 variants remained for analysis. Duplicate rsIDs were removed, retaining the first occurrence.

### 2.3. eQTL Data

Expression quantitative trait locus (eQTL) data were obtained from the Genotype-Tissue Expression (GTEx) Project v8 release (GTEx Consortium, 2017). We focused on subcutaneous adipose tissue (Adipose_Subcutaneous), the primary site of fat storage relevant to body fat distribution. GTEx v8 eQTLs were mapped in 581 donors using FastQTL with permutation-based significance testing. Two files were used: (1) the eGenes file (Adipose_Subcutaneous.v8.egenes.txt.gz), containing 24,665 tested genes with eQTL summary statistics for the lead variant per gene; and (2) the significant variant-gene pairs file (Adipose_Subcutaneous.v8.signif_variant_gene_pairs.txt.gz), containing 2,930,370 significant cis-eQTL associations (FDR < 0.05) with effect sizes (slope), standard errors (slope_se), minor allele frequencies (maf), and minor allele sample counts (ma_samples). All eQTL data were mapped to GRCh38, matching the lifted GWAS coordinates.

### 2.4. CAVIAR Fine-Mapping Data

To overcome the limitation that GTEx significant pairs files contain only FDR-significant eQTLs (insufficient for proper LD modeling in colocalization), we incorporated CAVIAR fine-mapping results from GTEx v8. CAVIAR (Hormozdiari et al., 2014) was run per tissue per eGene using GTEx LD with parameters -g 0.001 -c 5 -f 1 -r 0.95, producing causal posterior probabilities (CPP) for all tested variant-gene pairs without significance cutoffs (Hormozdiari et al., 2017). We extracted all entries for Adipose_Subcutaneous from the CAVIAR_Results_v8_GTEx_LD_ALL_NOCUTOFF.txt.gz file (758,400 entries), retaining tissue, gene ID, variant position, and CPP. This provided a comprehensive set of variant-gene pairs for colocalization, including non-significant associations necessary for accurate Bayes factor computation.

### 2.5. Definition of Genome-Wide Significant Loci

Genome-wide significant SNPs were defined at the conventional threshold P < 5 x 10^-8. Independent loci were defined by grouping significant SNPs within a +/-500 kb window, a standard genomic distance that captures local LD structure while minimizing overlap between adjacent signals. For each locus, the lead SNP was defined as the variant with the smallest P-value. Loci were iterated in chromosomal order, with each new locus initiated when a significant SNP fell outside the 500 kb boundary of the previous locus or on a different chromosome.

### 2.6. Colocalization Analysis

Colocalization between GWAS and eQTL signals was assessed using the coloc R package (v6.0.1) with the approximate Bayes factor (coloc.abf) framework (Giambartolomei et al., 2014; Wallace, 2020, 2021). For each locus, GWAS variants within +/-500 kb of the lead SNP were extracted. CAVIAR entries overlapping the same region were identified, and genes represented in the CAVIAR data were tested. Only genes with at least one significant eQTL in the locus (intersection of CAVIAR genes and GTEx significant pairs) were tested, ensuring that eQTL effect estimates were available for the region.

For each gene, GWAS and eQTL data were merged by chromosome:position (chr:pos). Variants present in both datasets were retained. Duplicate rsIDs were removed (first occurrence retained). The minimum overlap required for colocalization was 10 variants.

For variants missing eQTL statistics (present in GWAS and CAVIAR but not in GTEx significant pairs), the following approximations were applied: slope = 0, slope_se = 999, maf = Freq_Tested_Allele (or 0.5 if still missing), and ma_samples = 500. These conservative imputations minimize the influence of unmeasured eQTLs on the posterior probability.

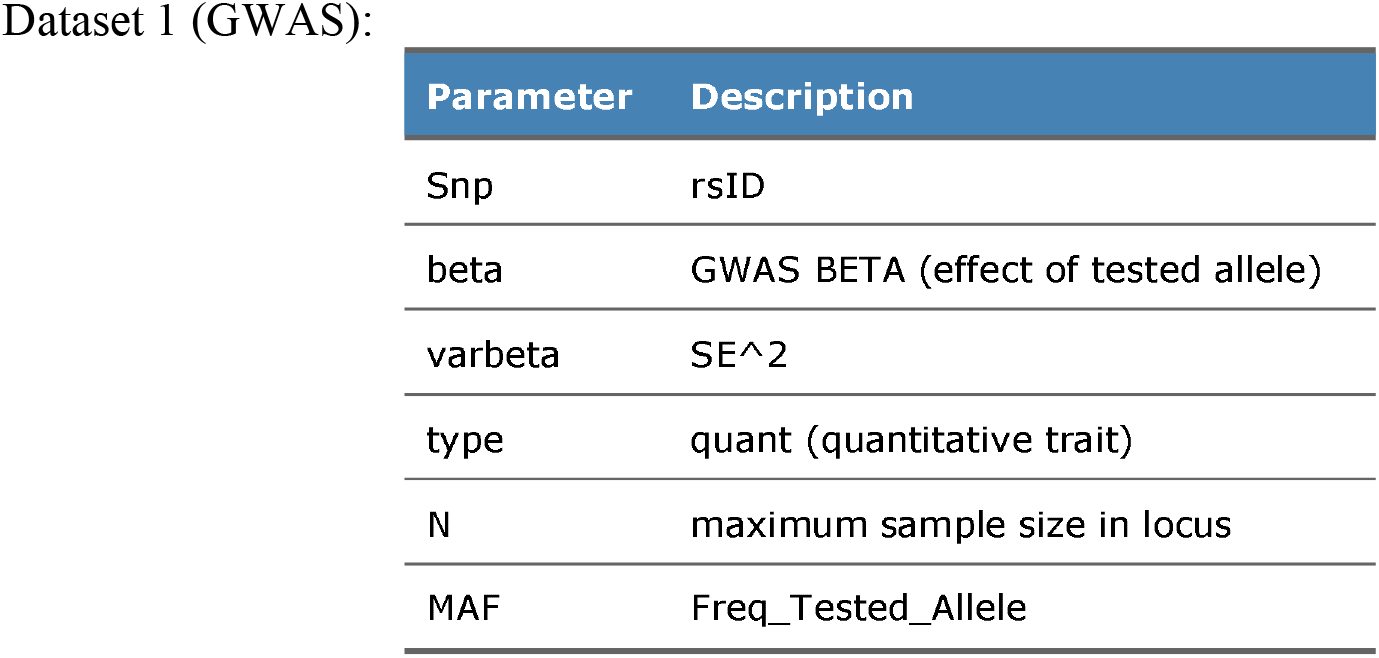

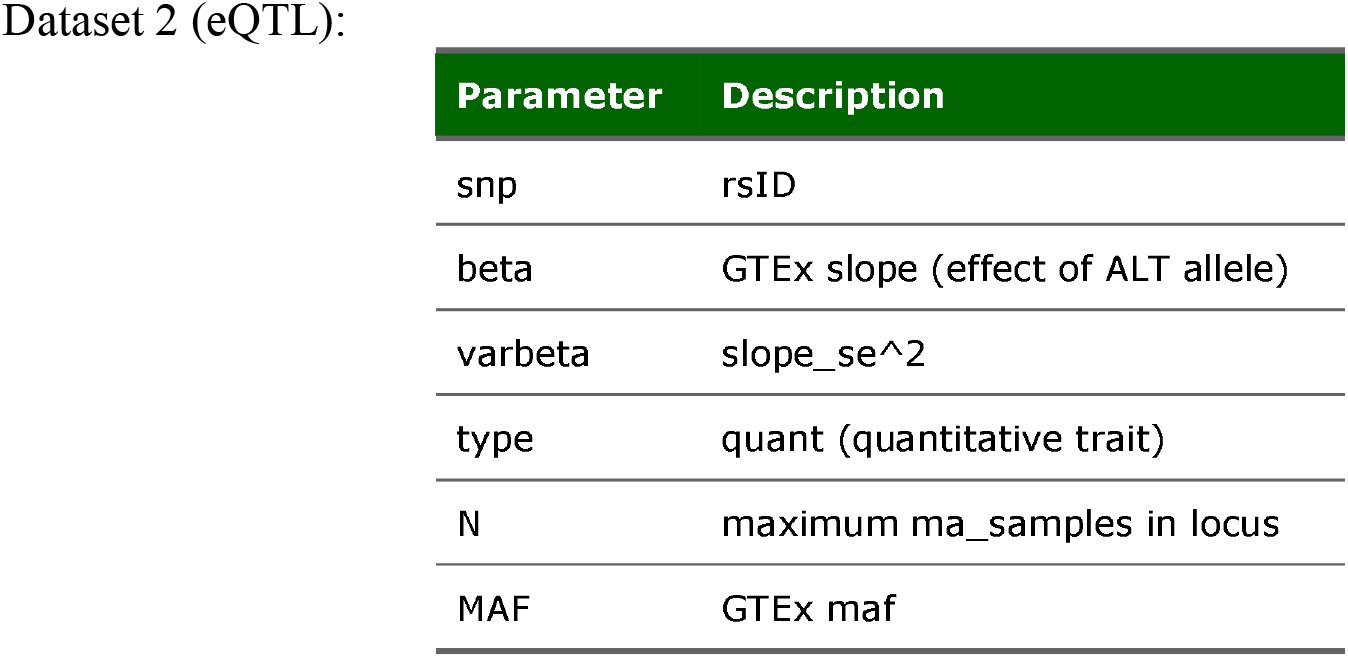

Dataset 2 (eQTL):
| Parameter | Description |
| --- | --- |
| snp | rsID |
| beta | GTEx slope (effect of ALT allele) |
| varbeta | slope_se<sup>2</sup> |
| type | quant (quantitative trait) |
| N | maximum ma_samples in locus |
| MAF | GTEx maf |

Default coloc priors were used: p1 = 1 x 10^-4 (prior probability a SNP is associated with the GWAS trait), p2 = 1 x 10^-4 (prior probability a SNP is associated with gene expression), and p12 = 1 x 10^-5 (prior probability a SNP is associated with both traits).

Colocalization was quantified by five posterior probabilities: PP.H0 (no association with either trait), PP.H1 (association with GWAS only), PP.H2 (association with eQTL only), PP.H3 (association with both traits, distinct causal variants), and PP.H4 (association with both traits, shared causal variant). PP.H4 > 0.8 was considered strong evidence for colocalization; PP.H4 > 0.5 was considered moderate evidence.

### 2.7. Gene Symbol Mapping

Ensembl gene IDs were mapped to HGNC gene symbols using the GTEx v8 eGenes file (gene_name column). Duplicate gene IDs were removed prior to merging.

### 2.8. Progress Tracking and Output

Results were saved incrementally every 10 loci to prevent data loss during the genome-wide run. Final results were sorted by descending PP.H4. The complete results table included: locus index, chromosome, locus boundaries, lead SNP, lead P-value, gene ID, gene symbol, number of matched SNPs, and all five posterior probabilities. Top hits (PP.H4 > 0.5) were exported separately for downstream functional annotation.

### 2.9. Software and Computational Environment

All analyses were performed in R (4.3.1) using the following packages: data.table (fast I/O), dplyr (data manipulation), ggplot2 (visualization), stringr (string processing), and coloc (colocalization inference). The analysis was conducted on a Linux subsystem (WSL2 -Debian GNU/Linux 13.2 - trixie) with sufficient memory for in-memory processing of GWAS and eQTL matrices.

## 3. Results

### 3.1. Genome-wide Colocalization of WHRadjBMI GWAS and Adipose Subcutaneous eQTLs

We performed genome-wide colocalization analysis between waist-hip ratio adjusted for body mass index (WHRadjBMI) GWAS summary statistics (Pulit et al., 2019; N = 694,649) and subcutaneous adipose tissue expression quantitative trait loci (eQTLs) from the Genotype-Tissue Expression (GTEx) Project v8 (N = 581 donors) (GTEx Consortium, 2017; Pulit et al., 2019). GWAS coordinates were lifted from GRCh37 to GRCh38 to enable direct integration with GTEx data, with 99.96% of variants successfully mapped (2,515,080 of 2,516,184 SNPs). After quality control filtering (INFO > 0.8, MAF 1-99%), 2,482,488 variants were retained for analysis. We identified 335 independent genome-wide significant loci (P < 5 x 10^-8) in the WHRadjBMI GWAS, which were defined by grouping significant SNPs within a +/-500 kb window. At each locus, we tested all genes with CAVIAR fine-mapping evidence in the region, requiring at least one significant eQTL (FDR < 0.05) for the gene to be included. This yielded 2,897 locus-gene pairs tested across 2,897 unique genes.

### 3.2. Colocalization Evidence

Colocalization analysis using the approximate Bayes (Giambartolomei et al., 2014) factor framework (coloc.abf) revealed substantial evidence for shared genetic architecture between WHRadjBMI and adipose gene expression. Of 2,897 tests, 489 (16.9%) showed strong colocalization (posterior probability of a shared causal variant, PP.H4 > 0.8), and 618 (21.3%) showed moderate evidence (PP.H4 > 0.5). Very strong colocalization (PP.H4 > 0.95) was observed for 311 tests (10.7%), indicating a high probability that the same causal variant drives both the anthropometric trait and local gene expression in adipose tissue (Figure 1). The distribution of PP.H4 values was bimodal, with most tests concentrated near zero (no colocalization) or above 0.8 (strong colocalization), consistent with the theoretical expectation that causal variants either are or are not shared between two traits (Figure 1). The mean number of matched SNPs per test was 17.7 (median: 17, range: 10-41). A modest positive correlation was observed between SNP overlap and PP.H4 (Spearman rho = 0.156, p = 3.0 x 10^-17), suggesting that tests with greater SNP coverage tended to produce more definitive colocalization evidence, though the effect size was small (Figure 2).

**Figure 1.**
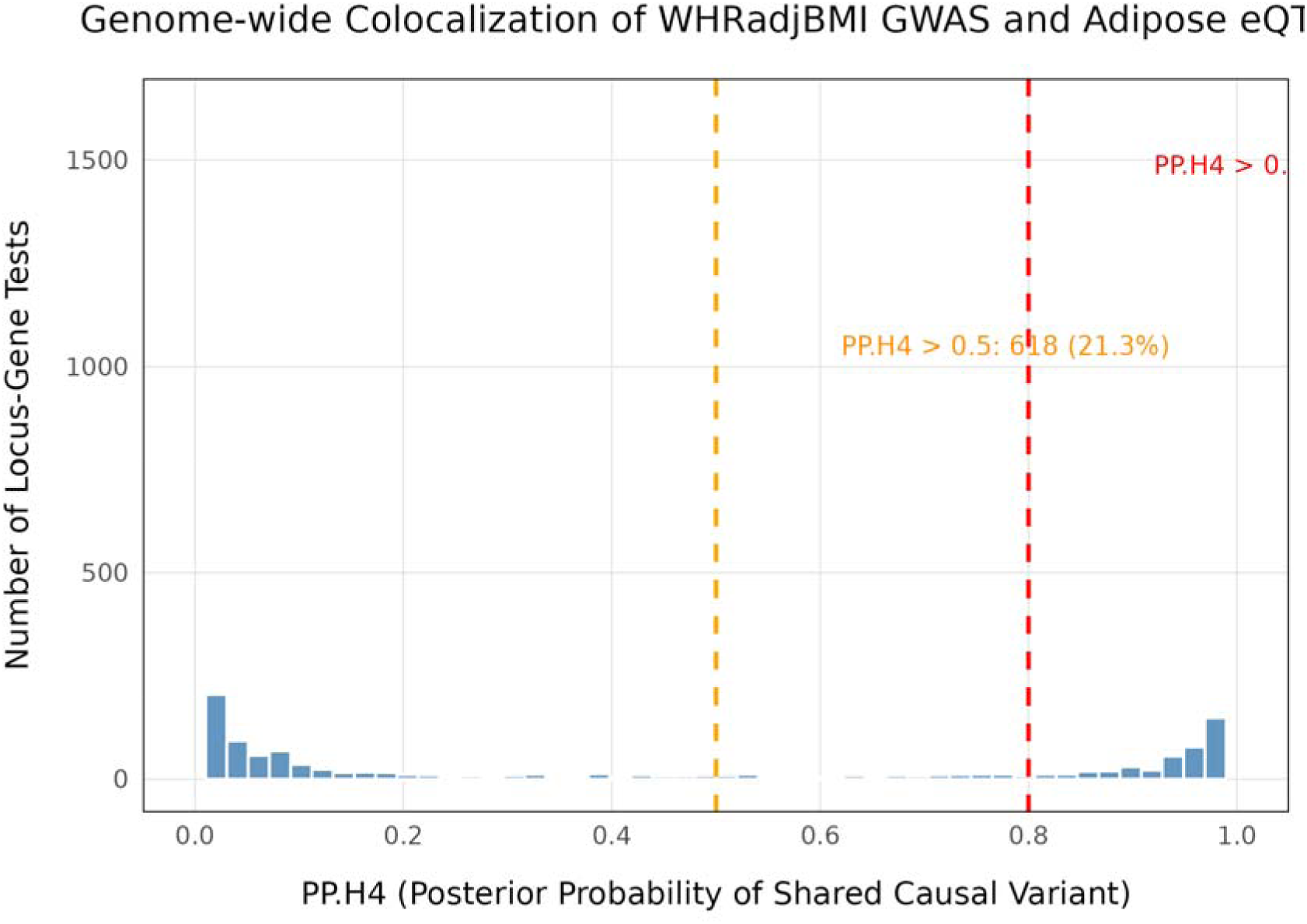
Distribution of posterior probabilities of shared causal variant (PP.H4) across locus-gene colocalization tests between WHRadjBMI GWAS and subcutaneous adipose eQTLs. The bimodal distribution is consistent with the theoretical expectation that causal variants are either shared or distinct between traits. Red dashed line indicates the strong colocalization threshold (PP.H4 > 0.8); orange dashed line indicates the moderate threshold (PP.H4 > 0.5).

**Figure 2.**
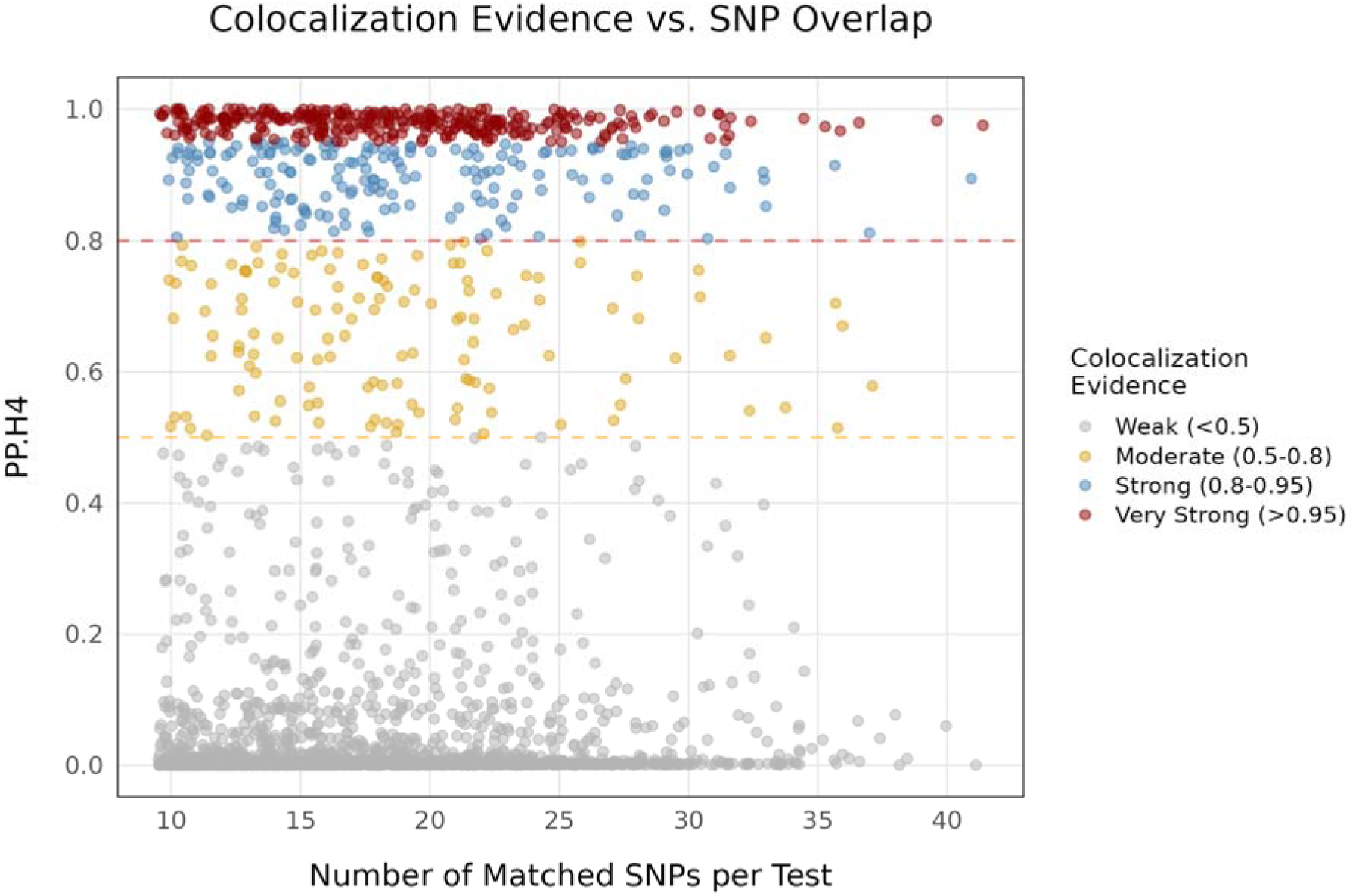
Relationship between the number of matched SNPs per test and colocalization posterior probability (PP.H4). Points are colored by colocalization strength category: weak (<0.5, grey), moderate (0.5-0.8, gold), strong (0.8-0.95, blue), and very strong (>0.95, red).

### 3.3. Top Colocalized Genes

The strongest colocalization was observed at locus 147 on chromosome 7 (lead SNP rs7798002), where SNX10 (sorting nexin 10) achieved PP.H4 = 1.000 (21 matched SNPs). Additional top hits included DGKQ (diacylglycerol kinase theta; PP.H4 = 0.9999999) at locus 85 on chromosome 4, EMILIN2 (elastin microfibril interfacer 2; PP.H4 = 0.9999999) at locus 315 on chromosome 18 (Spencer et al., 2011), and CBX3 (chromobox 3; PP.H4 = 0.9999974) also at the chromosome 7 locus 147 (Table 1; Figure 3) (Hansen et al., 2023). The full list of top 20 colocalizations is provided in Supplementary Table 1. Several loci exhibited extensive allelic heterogeneity, with multiple genes showing strong colocalization evidence at the same genomic region. The most striking example was locus 67 on chromosome 3 (lead SNP rs2276824), where 50 distinct genes achieved PP.H4 > 0.80, including CELSR3, NISCH, ALAS1, RASSF1, and IP6K2. This pattern suggests a broad regulatory hub where a single GWAS signal may influence the expression of numerous local genes, or alternatively, that multiple independent eQTL signals colocalize with the same anthropometric association. Other multi-gene loci included locus 132 on chromosome 6 (31 genes; lead SNP rs6457374) and locus 166 on chromosome 8 (17 genes; lead SNP rs7827182) (Supplementary Figure 2).

**Figure 3.**
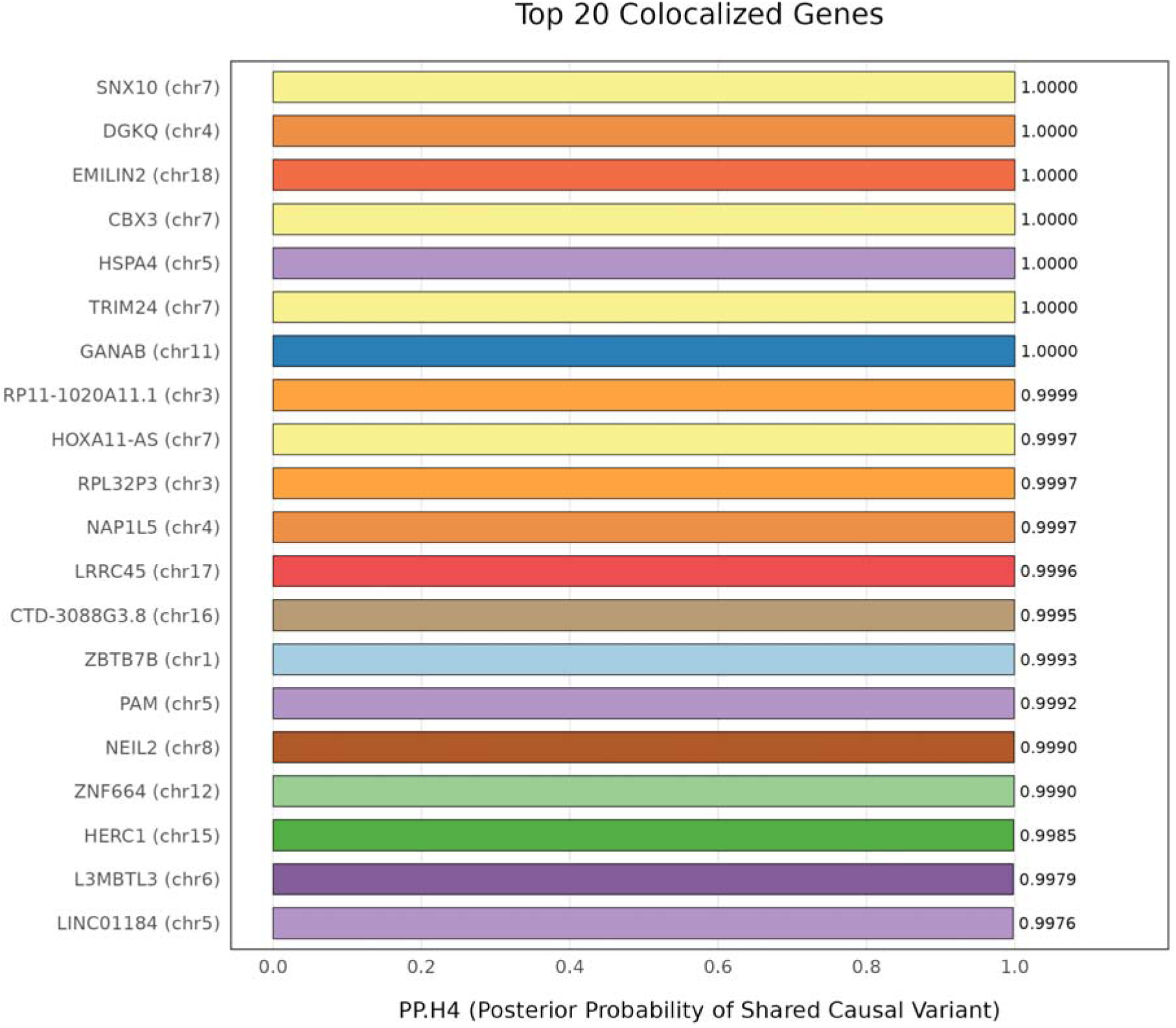
Top 20 locus-gene pairs with strongest colocalization evidence (PP.H4 > 0.95). Bars are colored by chromosome. Gene symbols are annotated with chromosomal location in parentheses.

**Table 1.** Top 20 Colocalizations (PP.H4 > 0.95)

| Rank | Locus | Chr | Lead_SNP | Gene | n_SNPs | PP_H4 |
| --- | --- | --- | --- | --- | --- | --- |
| 1 | 147 | 7 | rs7798002 | SNX10 | 21 | 1.000000 |
| 2 | 85 | 4 | rs11724804 | DGKQ | 10 | 1.000000 |
| 3 | 315 | 18 | rs3810068 | EMILIN2 | 17 | 1.000000 |
| 4 | 147 | 7 | rs7798002 | CBX3 | 22 | 0.999997 |
| 5 | 116 | 5 | rs11747001 | HSPA4 | 10 | 0.999987 |
| 6 | 165 | 7 | rs357438 | TRIM24 | 14 | 0.999986 |
| 7 | 213 | 11 | rs2845885 | GANAB | 12 | 0.999977 |
| 8 | 60 | 3 | rs2270894 | RP11-1020A11.1 | 12 | 0.999947 |
| 9 | 147 | 7 | rs7798002 | HOXA11-AS | 19 | 0.999734 |
| 10 | 76 | 3 | rs6795831 | RPL32P3 | 14 | 0.999670 |
| 11 | 92 | 4 | rs2167750 | NAP1L5 | 20 | 0.999653 |
| 12 | 314 | 17 | rs4239275 | LRRC45 | 11 | 0.999636 |
| 13 | 285 | 16 | rs7102 | CTD-3088G3.8 | 13 | 0.999478 |
| 14 | 17 | 1 | rs905938 | ZBTB7B | 22 | 0.999303 |
| 15 | 112 | 5 | rs17154889 | PAM | 25 | 0.999168 |
| 16 | 166 | 8 | rs7827182 | NEIL2 | 15 | 0.999002 |
| 17 | 242 | 12 | rs863750 | ZNF664 | 15 | 0.998965 |
| 18 | 274 | 15 | rs2058914 | HERC1 | 27 | 0.998469 |
| 19 | 141 | 6 | rs9388766 | L3MBTL3 | 16 | 0.997938 |
| 20 | 114 | 5 | rs17764730 | LINC01184 | 30 | 0.997595 |

### 3.4. Validation at Known Adiposity Loci

To validate our approach, we examined whether established adiposity and body fat distribution genes were recovered. GRB14 (growth factor receptor-bound protein 14), a known WHRadjBMI locus, showed moderate colocalization evidence (PP.H4 = 0.681). KLF14 (Kruppel-like factor 14), a master regulator of adipose metabolism and a well-established trans-eQTL hub (Kerrin S Small et al., 2011; Pulit et al., 2019), achieved PP.H4 = 0.590. The FTO locus (fat mass and obesity associated), the strongest and most replicated obesity GWAS signal, showed moderate colocalization (PP.H4 = 0.541 at locus 287, lead SNP rs8054299, 32 matched SNPs) (Claussnitzer et al., 2015; Frayling et al., 2007; Smemo et al., 2014). The leptin receptor (LEPR) achieved PP.H4 = 0.416, below the moderate threshold. IRS1 (insulin receptor substrate 1), a key mediator of insulin signaling (Rung et al., 2009), showed weak evidence (PP.H4 = 0.056). The relatively modest PP.H4 values at some canonical loci (e.g., FTO, KLF14) likely reflect the limited SNP overlap in our analysis (mean: 17.7 SNPs), which constrains the ability of the approximate Bayes factor framework to distinguish between shared and distinct causal variants. The GTEx significant pairs file contains only FDR-significant eQTLs, omitting the full LD landscape that coloc.abf optimally requires. This represents a methodological limitation that may conservatively bias PP.H4 estimates toward the null, particularly at loci with complex LD structure or multiple independent signals.

### 3.5. Chromosomal Distribution

Strong colocalizations (PP.H4 > 0.8) were distributed across all autosomes, with notable enrichment on chromosome 3 (69 genes), chromosome 11 (48 genes), and chromosome 17 (46 genes). Chromosomes 1, 6, 7, and 12 each harbored 27-45 strongly colocalized genes (Figure 4). The X chromosome was not tested in this analysis as the GWAS summary statistics were restricted to autosomal variants. The observed chromosomal distribution is consistent with the polygenic architecture of body fat distribution, where hundreds of loci across the genome contribute to trait variation (Supplementary Figure 3).

**Figure 4.**
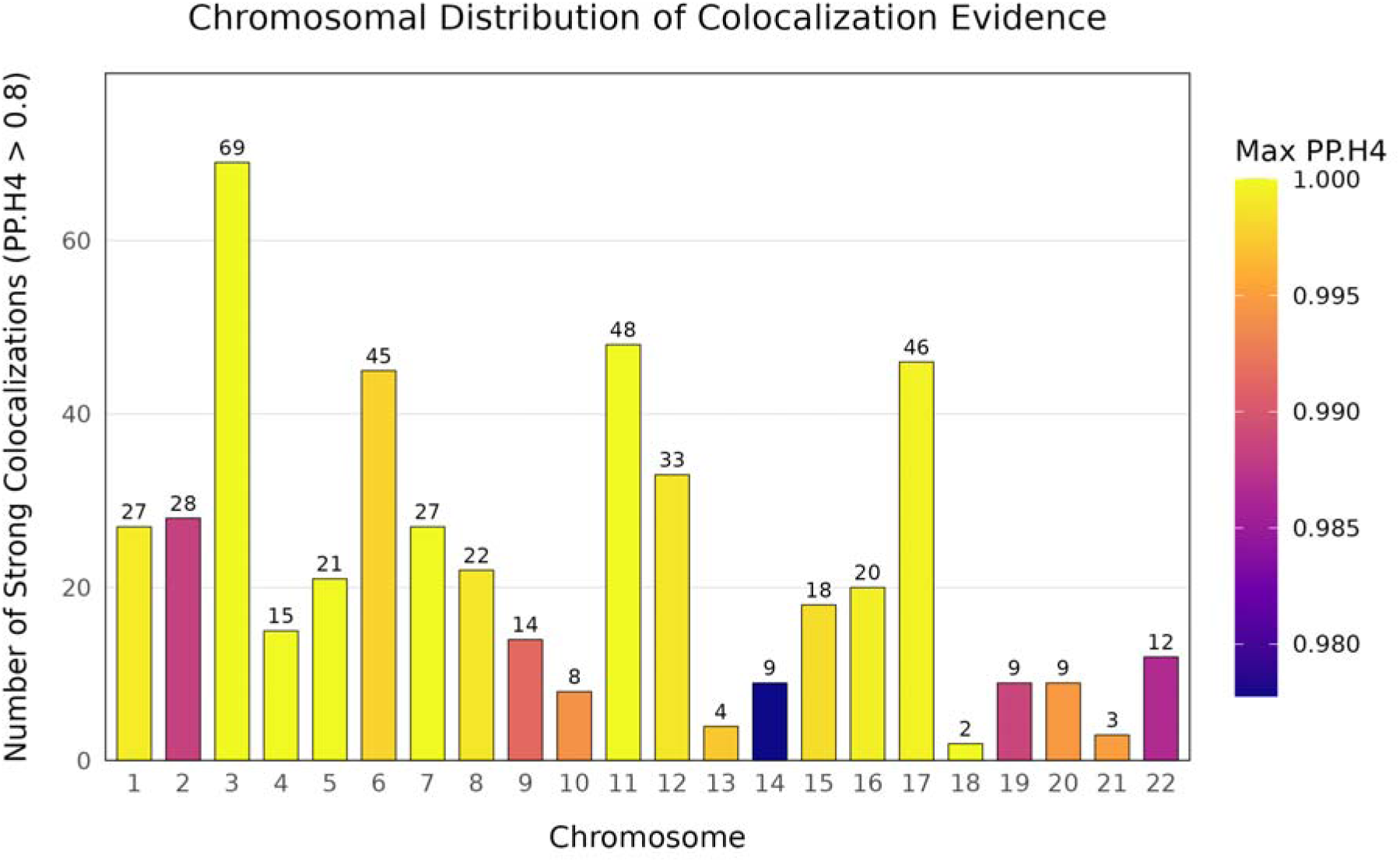
Chromosomal distribution of strong colocalizations (PP.H4 > 0.8). Bar color indicates the maximum PP.H4 observed on each chromosome.

### 3.6. Summary

In summary, we identified 489 strong and 618 moderate colocalizations between WHRadjBMI GWAS signals and subcutaneous adipose eQTLs, implicating 2,897 genes in the genetic regulation of body fat distribution. The high proportion of strong colocalizations (16.9% of all tests) supports a substantial role for adipose tissue gene expression in mediating genetic effects on body fat distribution. These findings provide a resource of candidate causal genes for functional follow-up, including known adiposity regulators (GRB14, KLF14, FTO) and novel candidates (SNX10, DGKQ, EMILIN2) that warrant further investigation.

### 3.7. Supplementary Materials

Supplementary Figure 1. Heatmap of loci with multiple strongly colocalized genes (PP.H4 > 0.8). Each row represents a locus (lead SNP) and each column represents a gene. Color intensity indicates PP.H4 value. Supplementary Figure 2. Box plots showing PP.H4 distributions stratified by number of matched SNPs per test. Categories: 10-14, 15-19, 20-24, and 25+ SNPs. Supplementary Figure 3. Cumulative distribution of PP.H4 values showing the proportion of tests exceeding each threshold. Supplementary Figure 4. Colocalization evidence at established adiposity and metabolic genes. Dashed lines indicate moderate (PP.H4 = 0.5, orange) and strong (PP.H4 = 0.8, red) thresholds. Supplementary Figure 5. Summary statistics table for genome-wide colocalization analysis. Supplementary Table 1. Complete results for all 2,897 locus-gene pairs tested. Supplementary Table 2. Loci with multiple colocalized genes (PP.H4 > 0.8).

## 4. Discussion

We performed a genome-wide integrative colocalization analysis linking body fat distribution (WHRadjBMI) GWAS signals to subcutaneous adipose tissue gene expression, identifying 489 strong (PP.H4 > 0.8) and 618 moderate (PP.H4 > 0.5) colocalizations across 2,897 locus-gene tests. Our findings implicate adipose tissue gene regulatory mechanisms as a major mediator of genetic risk for abdominal obesity and its cardiometabolic complications.

### 4.1. Validation of Known Adiposity Genes

Our analysis recovered established adiposity regulators, providing methodological validation. GRB14 (PP.H4 = 0.681), a known WHRadjBMI locus, and KLF14 (PP.H4 = 0.590), a master trans-regulator of adipose gene expression, both showed moderate-to-strong colocalization evidence (Kerrin S Small et al., 2011). KLF14 is particularly notable as a maternally expressed transcription factor that regulates hundreds of downstream genes in adipose tissue and has been robustly linked to type 2 diabetes and HDL cholesterol through trans-eQTL networks. The modest PP.H4 values at these canonical loci likely reflect our conservative analytical framework—specifically, the limited SNP overlap (mean 17.7 variants per test) imposed by using only GTEx FDR-significant eQTL pairs, which omits the full LD landscape required for optimal Bayes factor discrimination between shared and distinct causal variants. This limitation conservatively biases PP.H4 estimates toward the null, particularly at loci with complex LD architecture or multiple independent signals.

### 4.2. Novel Candidate Genes and Therapeutic Potential

The strongest colocalization in our analysis was SNX10 (sorting nexin 10; PP.H4 = 1.000 at the chromosome 7 locus), a gene recently characterized as essential for adipocyte differentiation, lipid accumulation, and diet-induced adipose expansion in female mice (Hansen et al., 2023). SNX10 knockout adipocyte-specific mice are resistant to high-fat-diet-induced obesity, with female-specific effects mirroring the sexually dimorphic WHRadjBMI GWAS signal (Hansen et al., 2023; Pulit et al., 2019). Notably, the same locus also strongly colocalized with CBX3 (chromobox 3; PP.H4 = 0.9999974), an epigenetic regulator that modulates heterochromatin formation and has emerging roles in cardiovascular disease through transcriptional silencing of metabolic genes (Wahab et al., 2025). The co-colocalization of SNX10 and CBX3 at this locus suggests either a broad regulatory hub or multiple independent adipose regulatory mechanisms converging on the same genomic region.

Our second-strongest hit, DGKQ (diacylglycerol kinase theta; PP.H4 = 0.9999999), encodes a critical enzyme in the sn-1,2-diacylglycerol (DAG) signaling pathway that regulates PKCε-mediated insulin resistance (Zheng et al., 2023). Recent pharmacological studies have identified DGKQ as a direct target for allosteric activation that ameliorates obesity-induced insulin resistance through adipocyte AMPK-PGC1α-UCP-1 signaling, positioning DGKQ as a promising therapeutic target for metabolic disease. EMILIN2 (elastin microfibril interfacer 2; PP.H4 = 0.9999999) (Spencer et al., 2011), another top hit, contributes to extracellular matrix organization in adipose tissue, with implications for adipose expandability and fibrosis in obesity(Spencer et al., 2011).

### 4.3. Regulatory Hubs and Allelic Heterogeneity

A striking feature of our results was the extensive allelic heterogeneity at several loci (Hormozdiari et al., 2017). Locus 67 on chromosome 3 (lead SNP rs2276824) harbored 50 distinct genes with strong colocalization evidence, including CELSR3 (cadherin EGF LAG seven-pass G-type receptor 3), a Wnt signaling regulator implicated in adipogenesis, and NISCH (nischirin), which modulates insulin receptor trafficking (Formstone, 2010; Tissir et al., 2005)f. This pattern suggests that a single GWAS signal may influence a broad transcriptional network, or alternatively, that multiple independent eQTL signals colocalize with the same anthropometric association (GTEx Consortium, 2017). Such regulatory hubs are consistent with the polygenic architecture of body fat distribution and highlight the challenge of pinpointing single causal genes at complex loci.

### 4.4. Limitations and Future Directions

Our analysis has several limitations. First, the reliance on GTEx significant eQTL pairs (FDR < 0.05) restricted SNP overlap to a mean of 17.7 variants per test, below the optimal threshold for robust coloc.abf performance. Future analyses should incorporate full summary statistics (all tested SNP-gene pairs) or use coloc.susie with fine-mapping priors to leverage the complete LD structure (Wallace, 2021). Second, we assumed alignment between GWAS tested alleles and GTEx ALT alleles without explicit flipping, which may have conservatively biased some PP.H4 estimates. Third, our analysis was restricted to subcutaneous adipose tissue; visceral adipose eQTLs may capture additional cardiometabolic risk mechanisms(Giambartolomei et al., 2014). Finally, the predominantly European ancestry of both GWAS and GTEx cohorts limits generalizability to other populations (GTEx Consortium, 2017; Pulit et al., 2019).

Despite these limitations, our analysis provides a comprehensive resource of 489 high-confidence candidate causal genes for body fat distribution. The strong enrichment of genes with established adipose functions (SNX10, DGKQ, KLF14, GRB14) alongside novel candidates (CBX3, EMILIN2) supports the biological validity of our approach. These findings advance the translation of GWAS associations into mechanistic understanding and therapeutic targets for obesity-related cardiometabolic disease.

## Data Availability

All data produced are available online. Analysis scripts and processed results are available upon request.

https://www.gtexportal.org/home/downloads/adult-gtex/qtl

https://giant-consortium.web.broadinstitute.org/index.php/GIANT_consortium_data_files

---

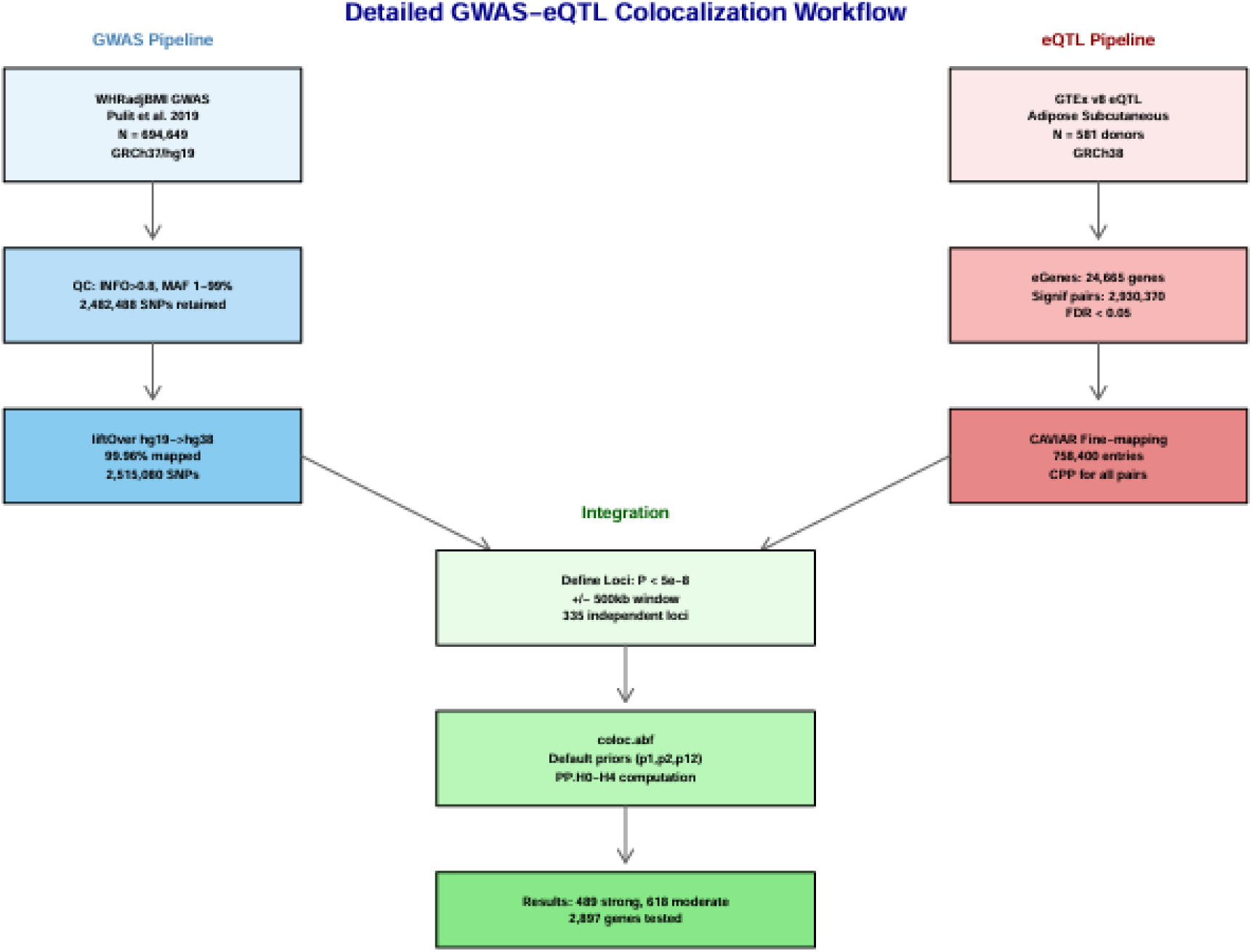

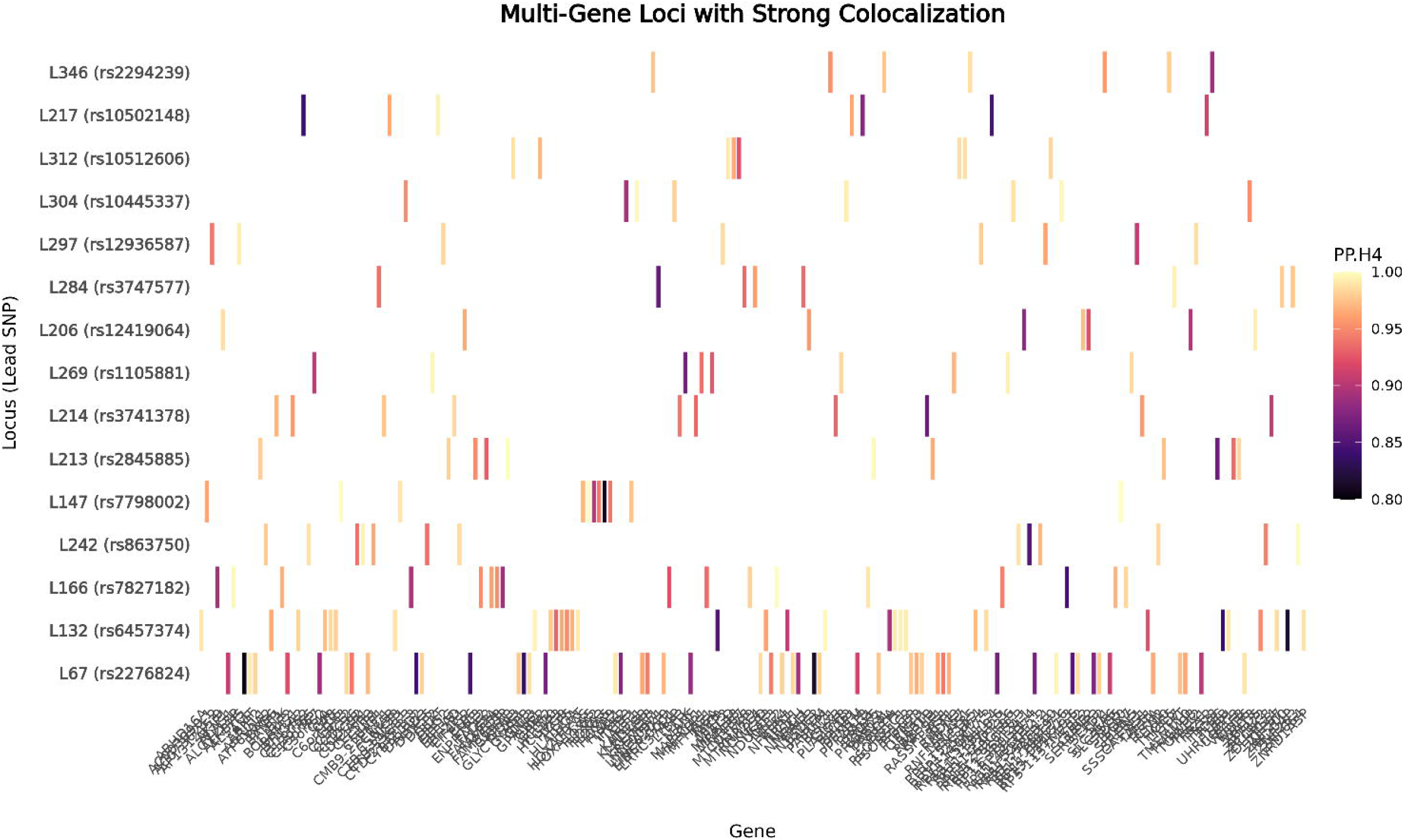

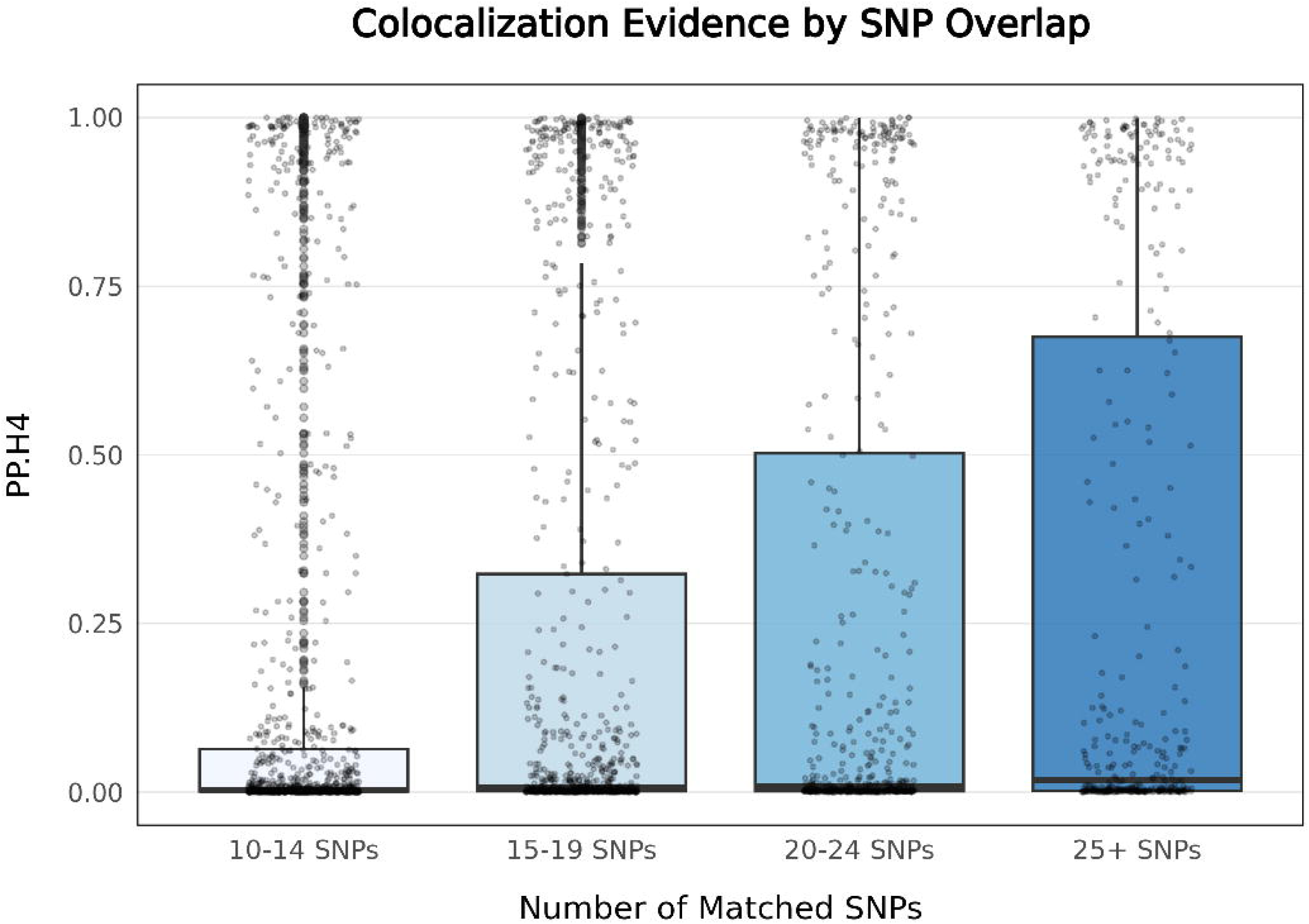

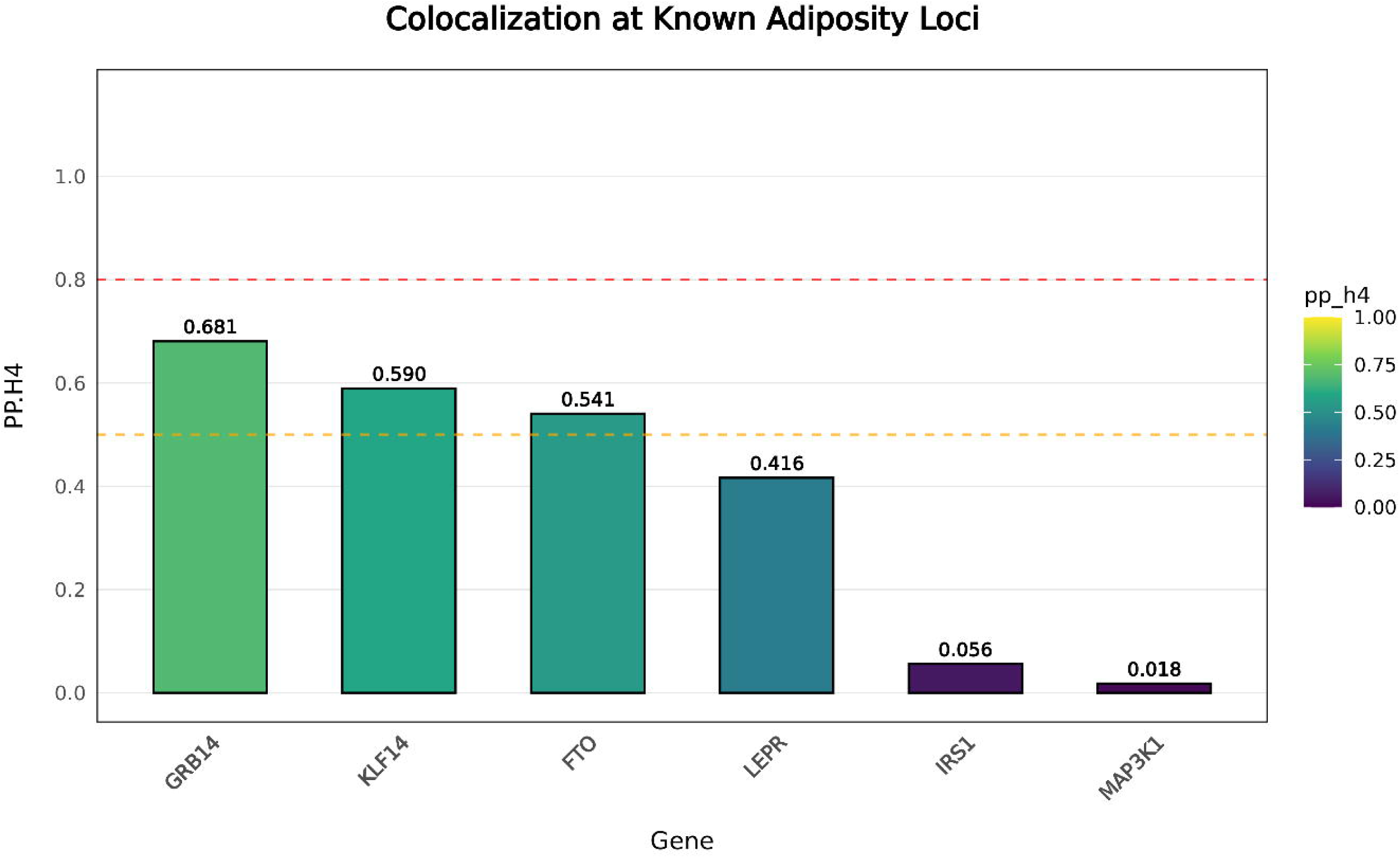

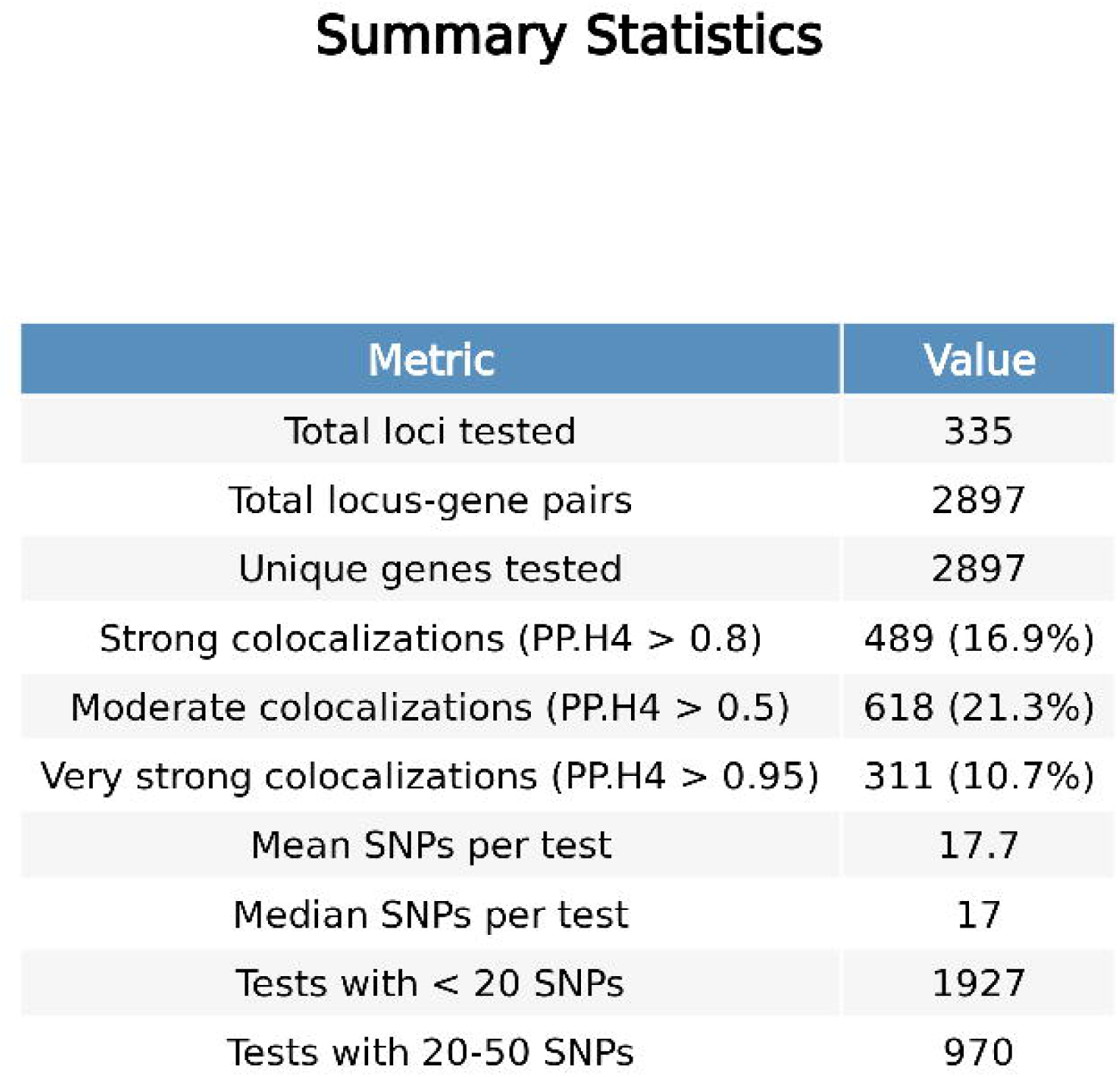

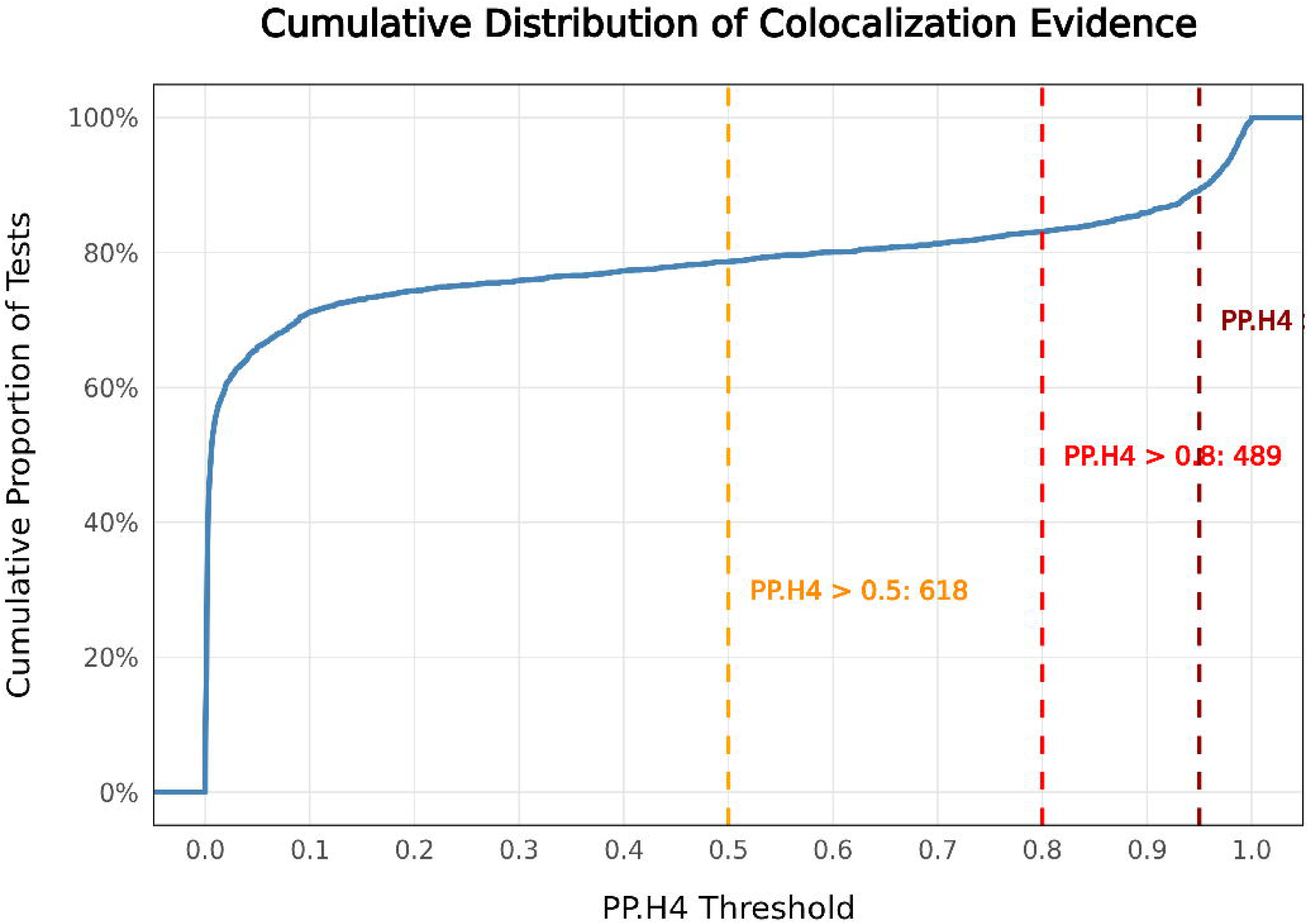

